# Prognostic Value of Different Iron Status Definitions in Congestive Heart Failure: A Retrospective MIMIC-IV Analysis of Risk Stratification and Mortality

**DOI:** 10.1101/2025.03.05.25323191

**Authors:** Abdulla Hourani, Arman David Sürmeli, Sai Keertana Devarapalli

## Abstract

**Background:** Iron deficiency (ID) is prevalent in congestive heart failure (CHF), worsening outcomes. While European guidelines recommend screening using ferritin and transferrin-saturation (TSAT), inconsistent diagnostic criteria, especially regarding functional deficiency (ferritin 100-299 µg/L + TSAT <20%) and hyperferritinemia, limit prognostic accuracy. This study evaluated iron status definitions, including guideline criteria and a combined Ferritin-TSAT model, for predicting 365-day mortality in hospitalized CHF patients.

**Methods:** This retrospective analysis used MIMIC-IV data from 1,839 CHF patients. Iron status within 24h of admission was categorized using: 1) Guideline ID vs. non-ID; 2) Ferritin categories; 3) TSAT categories; 4) Combined Ferritin-TSAT model (Low: guideline ID; Intermediate: ferritin 100-299 + TSAT ≥20%; High: ferritin ≥300 µg/L). Adjusted Cox models assessed mortality associations.

**Results:** Guidelines-defined iron deficiency (33.65% prevalence) independently associated with higher 1-year mortality (56.1% vs. 29.4%; adjusted HR 4.36, 95%CI 3.35-5.34). The combined Ferritin-TSAT model showed significant prognostic value, differentiating true iron deficiency (reference) from hyperferritinemia (adjusted HR 0.50 vs. iron deficiency) and intermediate group (adjusted HR 0.36 vs. ID), indicating varying risk relative to the most deficient group. This combined model better distinguished hyperferritinemic and iron-replete subgroups than the binary guideline definition.

**Conclusion:** Iron status, including deficiency and hyperferritinemia, independently predicts 1-year mortality in CHF. While guideline iron deficiency is a strong predictor, a combined Ferritin-TSAT classification offers finer risk stratification by identifying distinct phenotypes (true deficiency, hyperferritinemia, intermediate). Nuanced iron-status assessment could improve prognostic evaluation and guide personalized therapies (e.g., IV iron for deficiency, investigation for hyperferritinemia) to enhance CHF outcomes. Prospective validation is needed.

## Introduction

Iron deficiency anemia is a significant global public health concern and affects a substantial proportion of patients with congestive heart failure (CHF), with estimates suggesting that as many as 34–63% of individuals with chronic heart failure may be iron deficient [1]. Iron deficiency itself can adversely influence outcomes, by contributing to poor exercise tolerance, diminished health-related quality of life, and increased risks of hospitalization and mortality, even in the absence of anemia [2,3]. Recognizing the central role of iron deficiency in exacerbating congestive heart failure symptoms, the European Society of Cardiology has formally acknowledged iron deficiency as an important comorbidity and now recommends routine testing of iron status, including transferrin saturation (TSAT) and serum ferritin, for all newly diagnosed heart failure patients [4,5].

Iron deficiency in CHF patients can manifest as either absolute iron deficiency (depleted iron stores) or functional iron deficiency (adequate iron stores that cannot be effectively mobilized). Functional iron deficiency occurs when inflammatory processes in heart failure trigger increased production of hepcidin, which blocks iron release from storage sites despite normal or elevated ferritin levels. [6,7] This creates a state where iron is unavailable for critical cellular processes despite being present in the body [8,9]. The prevailing consensus holds that iron deficiency, irrespective of anemia, directly contributes to heart failure pathophysiology by impairing mitochondrial energy production, contributing to maladaptive cardiac remodeling, and activating inflammatory pathways that collectively reduce cardiac output [10].

Multiple large-scale trials and meta-analyses have consistently highlighted the clinical benefits of correcting iron deficiency in CHF. In particular, intravenous (IV) iron supplementation, most notably ferric carboxymaltose, has been shown to alleviate symptoms, improve exercise capacity, and reduce heart failure–related hospitalizations [4,11,12,13]. Observational and randomized evidence further indicate that these advantages extend beyond anemic patients to those with normal hemoglobin levels but low TSAT (<20%), which nonetheless signals depleted functional iron stores. IV iron administration improves left ventricular function indices, enhances functional status as measured by 6-minute walk tests, and reduces the rate of cardiovascular (CV) death or readmission for worsening congestive heart failure [11,14]. Oral iron supplementation, on the other hand, has generally not demonstrated consistent benefits in CHF, largely due to poor absorption and intolerability in this population [10]. This distinction reflects the necessity of targeting iron deficiency, guided by iron parameters like TSAT and ferritin, as a central therapeutic strategy in heart failure, rather than relying on hemoglobin levels to dictate treatment decisions.

However, the clinical utility of these findings is hindered by inconsistent diagnostic criteria, as there remains no universally accepted definition of iron deficiency in CHF for mortality prediction. Cutoff values for ferritin and TSAT vary among studies, with recent studies advocating for sole TSAT oriented operational definitions [15,16], creating a lack of standardization that complicates the analysis of iron deficiency prevalence, influencing patient therapy outcomes [1]. According to the most recent recommendations published by the European Society of Cardiology and their focused update, iron deficiency in heart failure is defined as a serum ferritin concentration <100 µg/L, or 100–299 µg/L with a transferrin saturation (TSAT) <20%, corresponding to absolute and functional iron deficiency, respectively [4,17,18]. Despite ESC guidance, the prognostic validity of these thresholds for long-term, post-discharge mortality remains unclear. Beyond classical deficiency, hyperferritinemia (≥300 μg/L), often reflecting inflammation or congestion, may be associated with a distinct risk profile; differentiating this phenotype from true deficiency may improve prognostic classification and motivate phenotype-specific evaluation.

In light of the above, this study focuses on further clarifying the effects and optimal diagnostic criteria in prognosis of iron deficiency in congestive heart failure. By examining the impact of iron deficiency on long-term mortality beyond the acute hospital setting, this research aims to address the ongoing need for clearer, evidence-based guidelines on screening and treating iron deficiency in CHF. To this end, the study rigorously evaluates and compares different stratification approaches, including a refined combined model, to overcome limitations in current definitions. Ultimately, these insights may contribute to improved quality of life and reduce the significant burden of recurrent hospitalizations and mortality among heart failure patients worldwide.

## Materials and Methods

### Study Design and Setting

We conducted a retrospective observational cohort study of adult hospitalizations with CHF using routinely collected electronic health record (EHR) data from a large tertiary academic medical center in the United States. The analysis was performed on the de-identified Medical Information Mart for Intensive Care IV (MIMIC-IV) database, which integrates information from intensive care units and affiliated hospital wards within Beth Israel Deaconess Medical Center; 2008-2019 [20]. The database contains detailed longitudinal data, including demographics, admission and discharge information, vital signs, laboratory measurements, procedures, diagnostic and procedure codes, medication information, and free-text clinical documentation for each hospitalization.

For this study, all eligible CHF inpatient stays recorded in the hospital module over the predefined study period were screened at the admission level and subsequently linked using unique patient identifiers to construct a cohort of index hospitalizations. Structured EHR tables (e.g., admissions, diagnoses, procedures, ICU stays, and laboratory results) were merged with unstructured clinical notes.

### Patient Selection

Inclusion criteria were ages between >18 and <89 years old, validated ICD-9/10 diagnosis of CHF, availability of admission notes, and iron parameters obtained within 24 hours of admission; only the first hospitalization per patient was retained. Patients with a length of stay <2 days were excluded, consistent with other studies. Patients aged ≥89 years were also excluded due to MIMIC age top-coding for data compliance, as this group is statistically rare and potentially re-identifiable.

Clinical notes were analyzed using a Gemma2-9B-Instruct–based extraction pipeline similar to the EchoLLM framework [19], but adapted to focus exclusively on left ventricular ejection fraction (LVEF). We prompted the model, with ten hand-crafted few-shot example echocardiography snippets, to return a structured JSON field containing LVEF as a numeric value (collapsing reported ranges to the most abnormal bound) and, when only qualitative language was present, to map LVEF into guideline-based categories (reduced, mildly reduced, preserved); few-shot prompting was used to enhance extraction accuracy [21,22]. Targeted regular-expression post-processing was applied to capture any remaining numeric EF mentions and to cross-check CHF diagnoses; and a random subset (n=100) of model outputs was manually compared with the source notes to validate correctness. Demographic and clinical variables (e.g., age, sex, comorbidities including shock, malignancy, renal disease, as well as ICU stay and ejection fraction status) were collected from structured EMR fields and supplemented by natural language processing. Laboratory measurements for ferritin, iron, TIBC, TSAT, hemoglobin, and other routine labs (e.g., creatinine for eGFR calculation) were obtained from the first tests within 24 hours of admission. Diagnoses were established via ICD-9/10 codes corroborated by notes. EF categories were used for phenotypic stratification and covariate adjustment. CKD was defined by ICD-9/10 codes for chronic renal insufficiency; AKI by ICD-9/10 codes consistent with KDIGO criteria. Procedures were identified using ICD procedure codes. Mineralocorticoid receptor antagonists (MRAs), angiotensin receptor–neprilysin inhibitors (ARNIs), and sodium–glucose cotransporter-2 inhibitors (SGLT2i) were not uniformly available or adopted in clinical practice during the study period. These therapies were not included in the present analysis.

### Definition of Iron Deficiency

Iron deficiency was defined using guideline-based thresholds of ferritin <100 μg/L or ferritin 100–299 μg/L with TSAT <20% [17]; non-iron deficiency otherwise. Analyses evaluated iron deficiency irrespective of anemia status; hemoglobin was included as an adjusted covariate and summarized at baseline.

### Data Cleaning and Categorization

All continuous variables were subjected to thorough assessment of distributional assumptions, including the Anderson–Darling test (for deviations in skewness and kurtosis), the Kolmogorov–Smirnov test (for overall distributional fit), and visual evaluation using histograms and Q–Q plots. Little’s test did not reject the hypothesis that data were missing completely at random (MCAR), and the observed missingness pattern was consistent with MCAR. Consequently, we proceeded with complete-case analysis.

Patients were categorized based on guideline-defined iron deficiency versus non-iron deficiency, clinically defined ferritin category, TSAT category, and a combined Ferritin-TSAT category classification. Given this study’s focus on mortality prediction, evaluating the prognostic utility of iron deficiency definitions was prioritized over distinguishing between functional and absolute subtypes, as clinical guidelines and prior evidence emphasize iron parameters’ collective role in risk stratification [9]. Category separation for the tested models was based on current guidelines and clinical thresholds. The Guidelines-based Model classified patients as iron deficient (ferritin <100 μg/L or ferritin 100–299 μg/L with transferrin saturation (TSAT) <20%) versus non–iron deficient. The Ferritin Category Model defined ferritin <100 μg/L as deficient, 100–299 μg/L as intermediate, and ≥300 μg/L as hyperferritinemic. In the TSAT Category Model, TSAT <10% was considered very low, 10–20% low, and >20% normal [23]. The combined Ferritin–TSAT Category Model defined low iron status as ferritin <100 μg/L or 100–299 μg/L with TSAT <20%, iron-replete as ferritin 100–299 μg/L with TSAT ≥20%, and hyperferritinemic as ≥300 μg/L. These classification frameworks were used to explore how variations in iron parameters relate to clinical outcomes while considering iron deficiency and overload.

### Statistical Analyses

Statistical analyses were conducted to compare differences across guideline-based iron deficiency groups, with the choice of parametric (t-tests, ANOVA) or nonparametric (Wilcoxon, Kruskal–Wallis) tests determined by distributional assessments. Categorical variables were compared using chi-square tests or Fisher’s exact test, depending on expected cell counts. Summary statistics for continuous variables were reported as means with standard deviations (SD) for normally distributed data or medians with interquartile ranges (IQR) for skewed distributions, while categorical variables were expressed as frequencies and percentages.

For time-to-event analysis, Kaplan–Meier survival curves were generated to visualize differences in 365-day mortality across study groups. Follow-up started at index admission and continued to 365 days, while only mortality was captured beyond 2019. Log-rank tests were performed to assess statistical significance between survival curves. To further quantify the association between iron parameters and mortality, Cox proportional hazards models were employed to estimate hazard ratios (HRs) with 95% confidence intervals (CIs). The proportional hazards assumption was evaluated using Schoenfeld residuals.

To account for potential confounders, three progressively adjusted Cox models were constructed for each classification framework. Model 1 adjusted for age, sex, ICU admission, and ejection fraction category, accounting for basic demographic and clinical severity factors. Model 2 built upon Model 1 by additionally adjusting for cardiac arrest, hypertension, diabetes, shock, malignancy, hemoglobin, and estimated glomerular filtration rate (eGFR) to incorporate comorbid conditions and laboratory markers relevant to both iron metabolism and cardiovascular outcomes. Model 3 further adjusted for dialysis, percutaneous coronary intervention (PCI) or coronary artery bypass grafting (CABG), beta blockers, angiotensin-converting enzyme inhibitors (ACEi) or angiotensin receptor blockers (ARBs), and iron supplementation, allowing for a comprehensive evaluation of treatment-related influences on mortality.

Subgroup analyses were performed on categories of: age ≥65 versus <65, sex, and reduced vs mildly-reduced vs. preserved ejection fraction to assess additional adjustments for potential confounding variables. Statistical significance was set at p < 0.05, and all analyses were performed using R (version 4.3).

### Ethical Considerations

This study was conducted in accordance with the principles of the Declaration of Helsinki and relevant institutional regulations for research using human data. All analyses were performed on the de-identified MIMIC-IV v2.2 database, which was created with approval from the institutional review boards of Beth Israel Deaconess Medical Center and the Massachusetts Institute of Technology, with a waiver of individual informed consent due to the use of de-identified data. Investigators completed the required training and data use agreements prior to accessing the database. Because the present work is a secondary analysis of an existing de-identified dataset, it did not involve any direct patient contact, did not affect clinical care, and posed minimal risk to participants. No attempt was made to re-identify individuals, and all data handling and reporting followed best practices to preserve patient privacy and confidentiality.

## Results

A total of 1,839 unique index admissions, with confirmed congestive heart failure, fulfilled all prespecified eligibility criteria; of which, 1220 had no guideline-based iron-deficiency, while 619 did meet the criteria. Marked imbalances in baseline characteristics were observed between iron deficient and non-iron deficient patients, and are summarized in Table 1. Iron deficient patients were older (median 77.0 [IQR 18.0] vs. 72.0 [18.0] years) and more frequently female (49.6% vs. 35.3%) (both p < 0.001). Laboratory contrasts included markedly lower ferritin (median 71.0 vs. 392.50 µg/L), iron (31.00 vs. 45.00 µg/dL) and TSAT (9.6% vs. 19.9%), but higher TIBC and transferrin (all p < 0.001). Clinically, they had fewer admissions for surgical interventions, shorter hospital stays and lower prevalence of shock (15% vs. 25%), acute kidney injury (14% vs. 26%), and myocardial infarction (14% vs. 23%), yet higher rates of atrial fibrillation (53% vs. 46%) and valvopathies (49% vs. 36%). Iron deficient patients had higher anticoagulant (51.4% vs. 40.1%) but lower vasopressor (20.8% vs. 36.1%), and ACEi/ARB exposure (28.8% vs. 32.5%) (all p < 0.001 except ACEi/ARB: p = 0.109). Despite higher 365-day mortality, the iron deficiency cohort had fewer surgical admissions (4.5% vs. 11.3%) and less ICU care (34.1% vs. 57.2%), with lower acute complications (AKI, MI, shock).

**Table 1.**
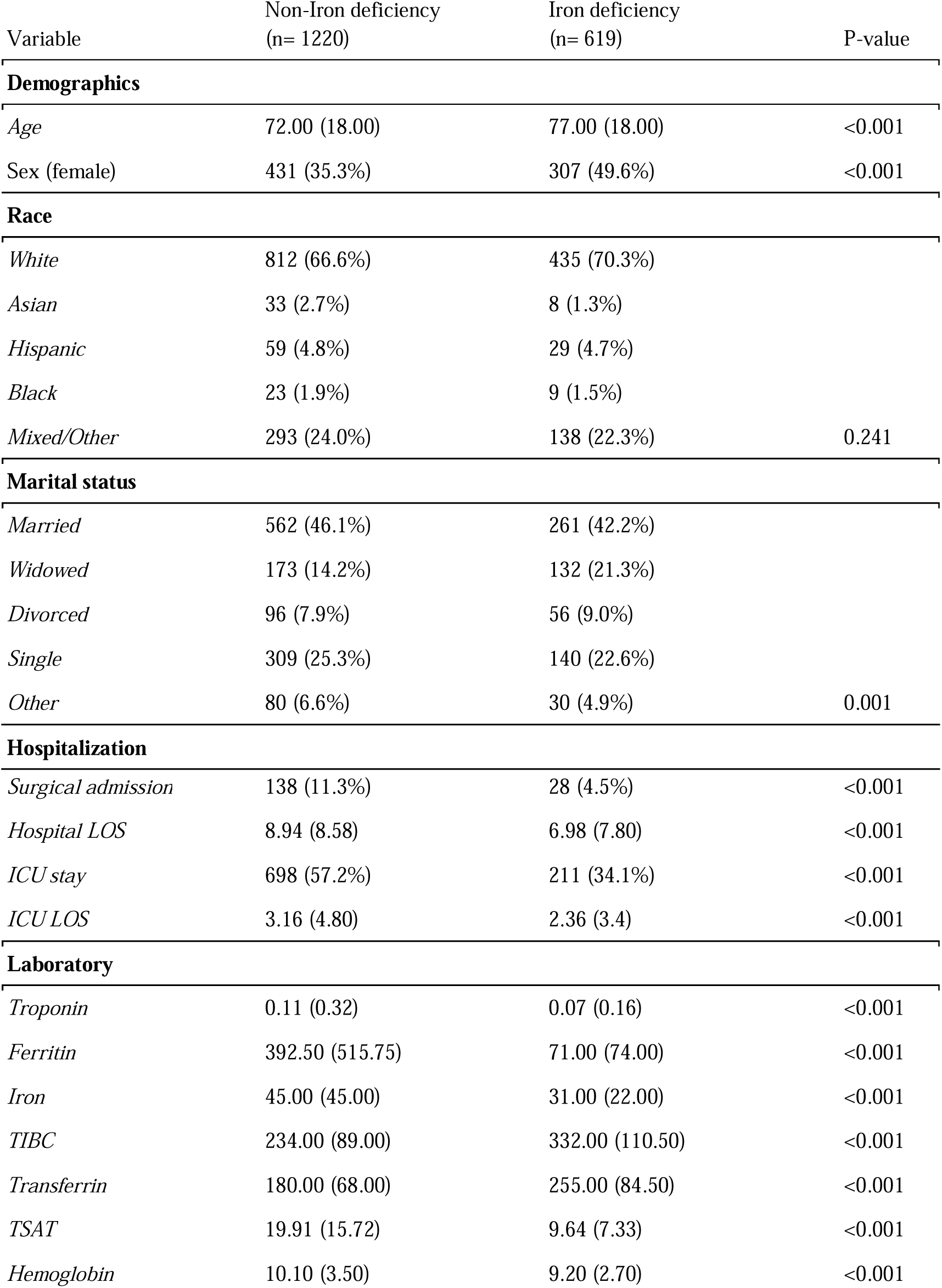
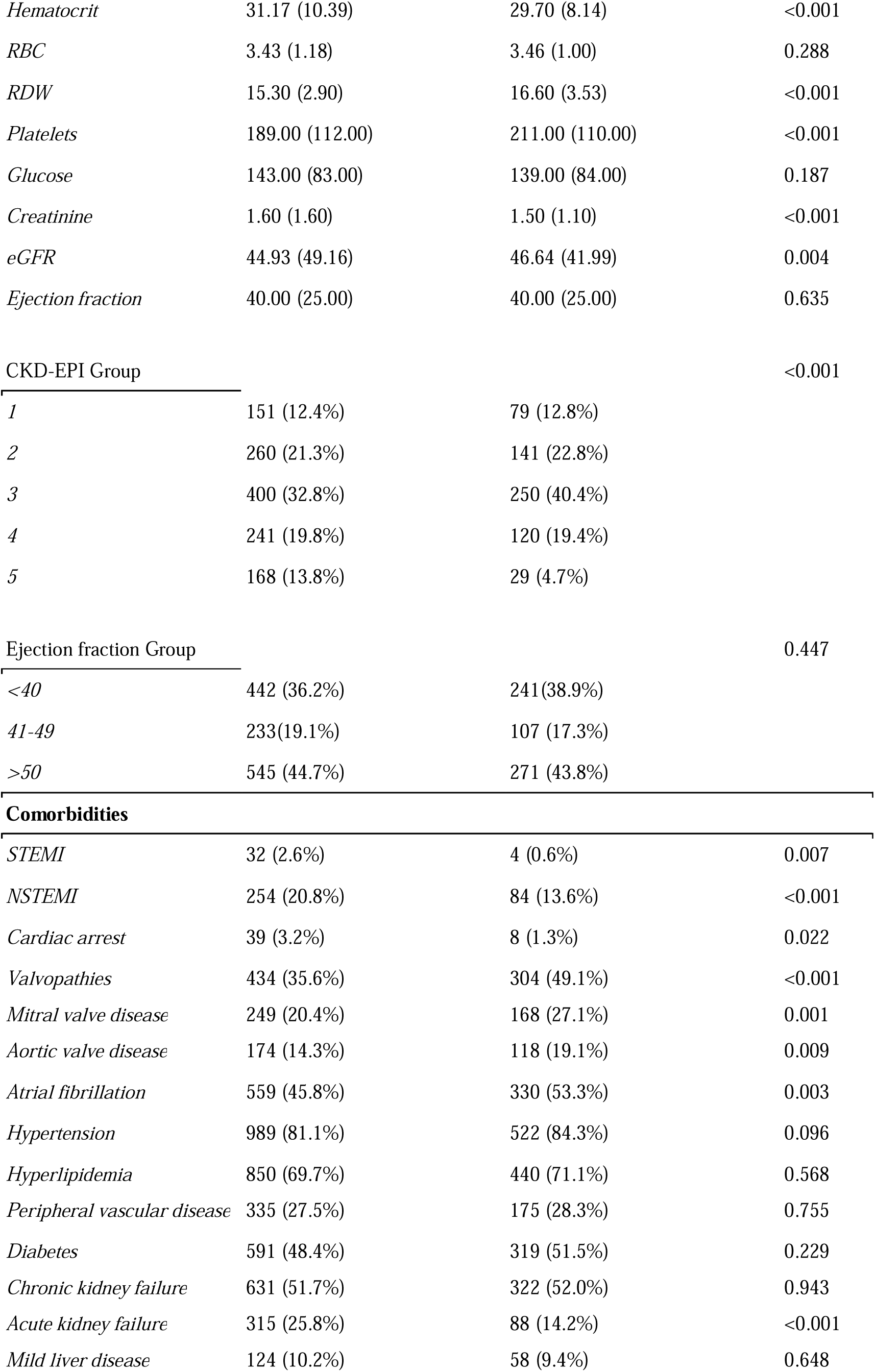
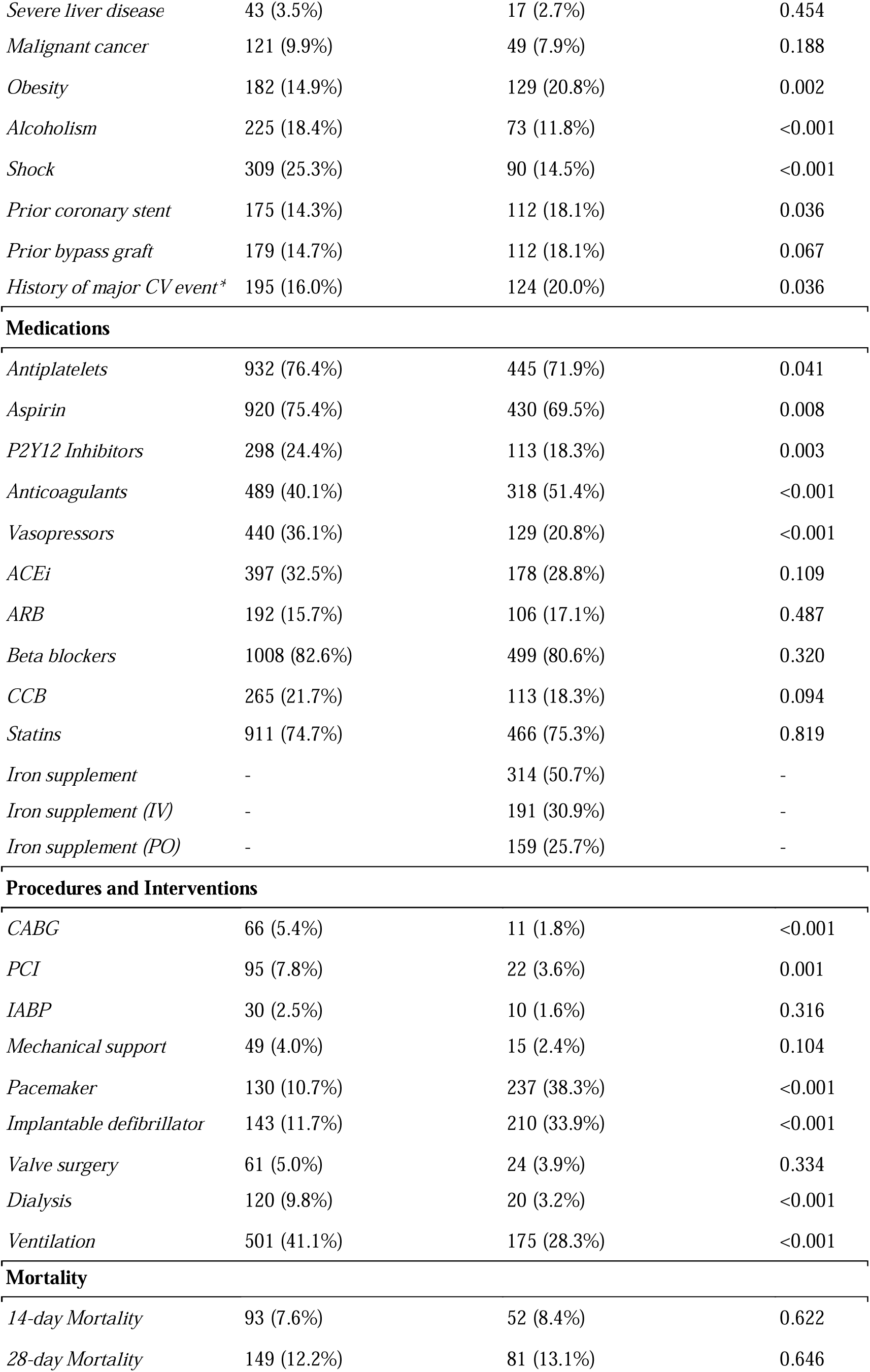
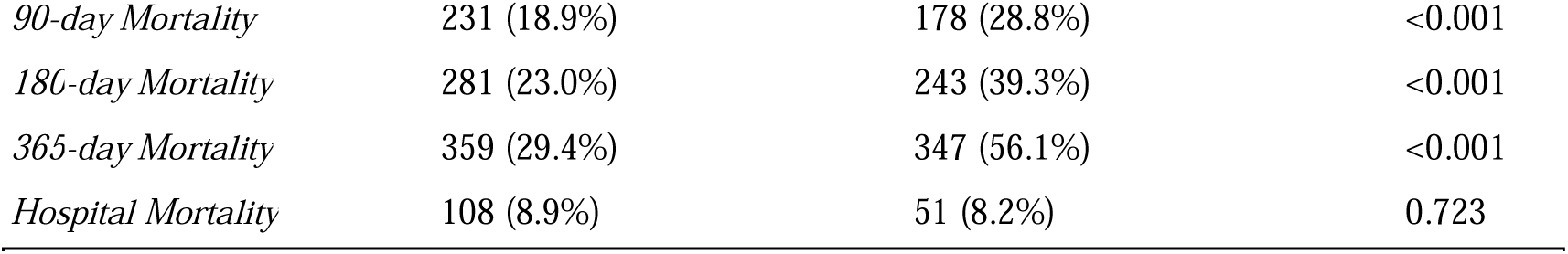
Baseline patient demographics, laboratory values, comorbidities, medications, procedures, and mortality outcomes stratified by iron deficiency status. Iron deficiency defined as ferritin <100 µg/L or ferritin 100–299 µg/L with TSAT <20% (guideline definition). Continuous variables are median (IQR). * History of CV outcomes constituted. Abbreviations: ACEi, angiotensin-converting enzyme inhibitor; ARB, angiotensin II receptor blocker; CABG, coronary artery bypass grafting; CCB, calcium channel blocker; CKD-EPI, Chronic Kidney Disease Epidemiology Collaboration; CV, cardiovascular; eGFR, estimated glomerular filtration rate; IABP, intra-aortic balloon pump; ICU, intensive care unit; IV, intravenous; LOS, length of stay; NSTEMI, non-ST-elevation myocardial infarction; PCI, percutaneous coronary intervention; PO, per os (oral); RBC, red blood cell count; RDW, red cell distribution width; STEMI, ST-elevation myocardial infarction; TIBC, total iron-binding capacity; TSAT, transferrin saturation.

Within this cohort, guideline-defined iron deficiency was present in 33.65% (N = 619), leaving 66.34% categorized as non-iron deficient. Application of the 3-tiered combined Ferritin–TSAT Category Model onto the same cohort reclassified patients into true iron deficiency (50.4%; ferritin <100 μg/L or ferritin 100–299 μg/L with TSAT <20%), hyperferritinemia (40.4%; ferritin ≥300 μg/L), and iron-replete (9.2%; ferritin 100–299 μg/L with TSAT ≥20%) groups, illustrating appreciable overlap yet clear segregation between the binary and composite definitions. Overall out-of-hospital 365Lday mortality was 38.4% (N = 706) with survival diverging sharply by iron status. GuidelineLdefined iron deficiency was associated with a crude oneLyear mortality of 56.1% compared with 29.4% among nonLdeficient patients (p < 0.001).

Throughout the four definition models, iron status retained a clear, independent association with 365-day mortality. Unadjusted survival analyses confirmed poorer outcomes with iron deficiency regardless of definition, with Kaplan–Meier curves illustrating survival for each definition displayed in Figures 1–4. Univariate analyses confirmed a strong crude, more than double mortality hazard association between guideline-defined iron deficiency and 365-day mortality (HR 2.48, 95%CI 2.09–2.94), whereas graded indices showed stepwise protection with rising ferritin (Ferritin Category Model: HR 0.49, 95%CI 0.39–0.60), or TSAT (TSAT Category Model: HR 0.72, 95%CI 0.65–0.81), or both (Ferritin-TSAT Category Model: HR 0.70, 95%CI 0.63–0.78) (Table 2). Relative to TSAT <10%, TSAT ≥20% associated with lower hazard (HR 0.63, 95%CI 0.49–0.81). In the Ferritin Category Model, ≥300 μg/L associated with lower hazard (HR 0.31, 95%CI 0.23–0.40). In the combined Ferritin-TSAT Category Model, the iron-replete and hyperferritinemia groups had HRs 0.36 (95%CI 0.24–0.53) and 0.50 (95%CI 0.38–0.51) vs. the deficient reference. After sequential, fully adjusted multivariate regression modeling for demographics, comorbidity and treatment, including age, sex, ICU stay, shock, malignancy, diabetes, renal function, hemoglobin, dialysis status, invasive procedures, and guidelineLdirected therapies (full list of covariates available in Supplementary Materials), these trends continued, as the Guidelines-based Model remained independently predictive with a 4.36-fold increase in 365-day mortality risk (95%CI 3.35–5.34), but lacked the granularity as seen in the Ferritin–TSAT category model, isolating hyperferritinemic and true non-iron deficiency phenotypes from the iron-deficient reference with HR 0.50 (95%CI 0.38–0.51) and HR 0.36 (95%CI 0.24–0.53) respectively. To note, in these category models, HR < 1 indicates reduced mortality risk relative to the most iron-deficient group (reference), not absolute treatment benefit (Table 2). Full covariate estimates for all fully adjusted models are shown in Tables S1–S4. Across frameworks, higher age, ICU stay, shock, diabetes and malignancy were associated with increased 365-day mortality, whereas CABG, ACEi/ARB therapy, β-blocker use and intravenous iron were each associated with lower hazard.

**Figure 1.**
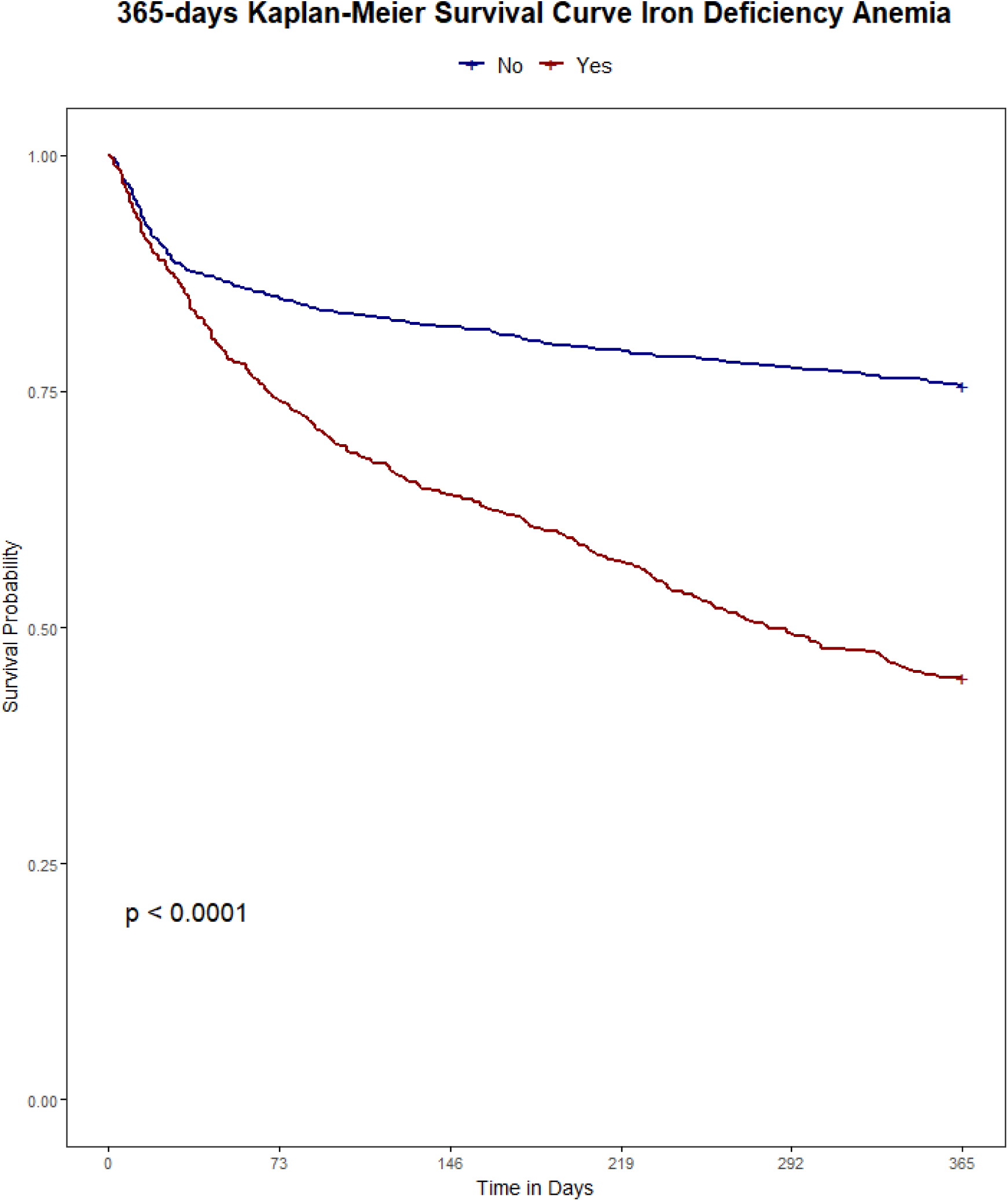
Kaplan–Meier survival to 365 days based on current guideline definitions. Population: hospitalized CHF index admissions. Groups: iron deficiency (ID) (ferritin <100 μg/L or ferritin 100–299 μg/L with transferrin saturation (TSAT) <20%) vs. non-ID. Time zero: admission. Outcome: all-cause mortality within 365 days. Deaths were ascertained from linked death records and counted within 365 days of admission, including events occurring after 2019; non-mortality follow-up was restricted to the dataset timeframe. Statistics: log-rank test with numbers at risk shown.

**Figure 2.**
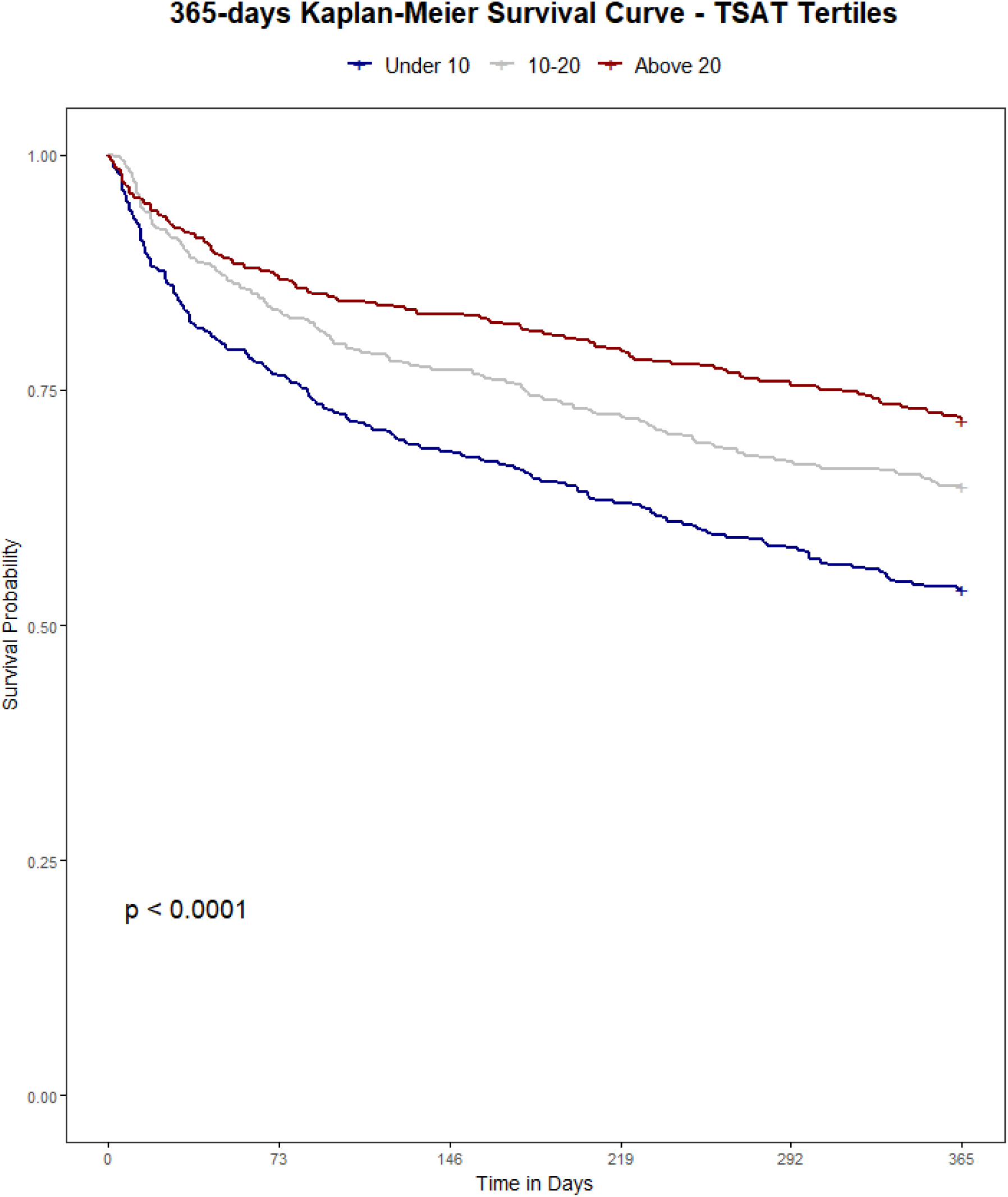
Kaplan–Meier survival to 365 days based on a TSAT category model. Population: hospitalized CHF index admissions. Groups: TSAT <10% (very low), TSAT 10–20% (low), TSAT ≥20% (normal). Time zero: admission. Outcome: all-cause mortality to day 365. Deaths were ascertained from linked death records and counted within 365 days of admission, including events occurring after 2019; non-mortality follow-up was restricted to the dataset timeframe. Statistics: log-rank test with numbers at risk shown. Abbreviations: TSAT, transferrin saturation.

**Figure 3.**
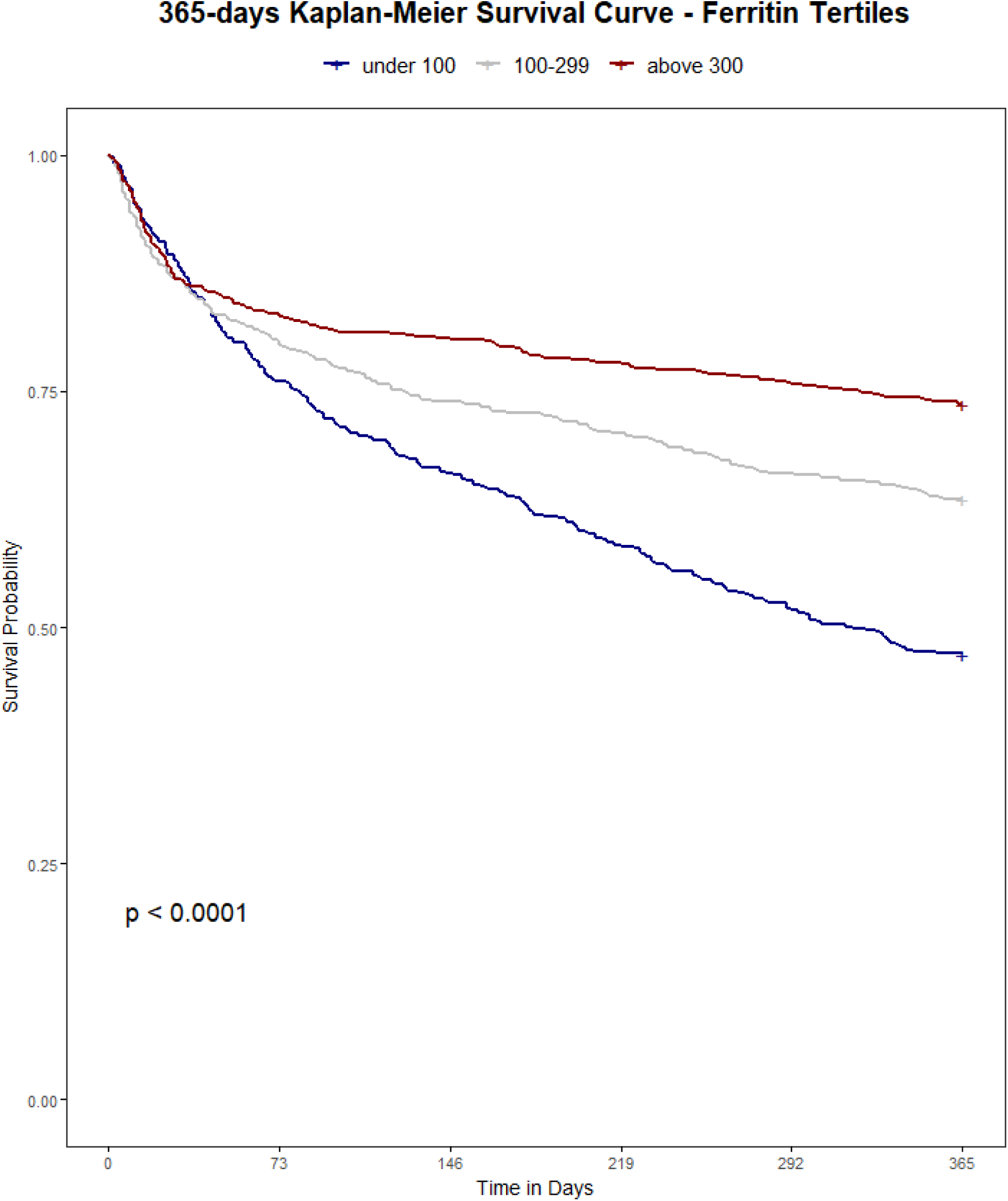
Kaplan–Meier survival to 365 days based on a Ferritin category model. Population: hospitalized CHF index admissions. Groups: ferritin <100 μg/L (low), 100–299 μg/L (intermediate), ≥300 μg/L (hyperferritinemia). Time zero: admission. Outcome: all-cause mortality to day 365. Deaths were ascertained from linked death records and counted within 365 days of admission, including events occurring after 2019; non-mortality follow-up was restricted to the dataset timeframe. Statistics: log-rank test with numbers at risk shown.

**Figure 4.**
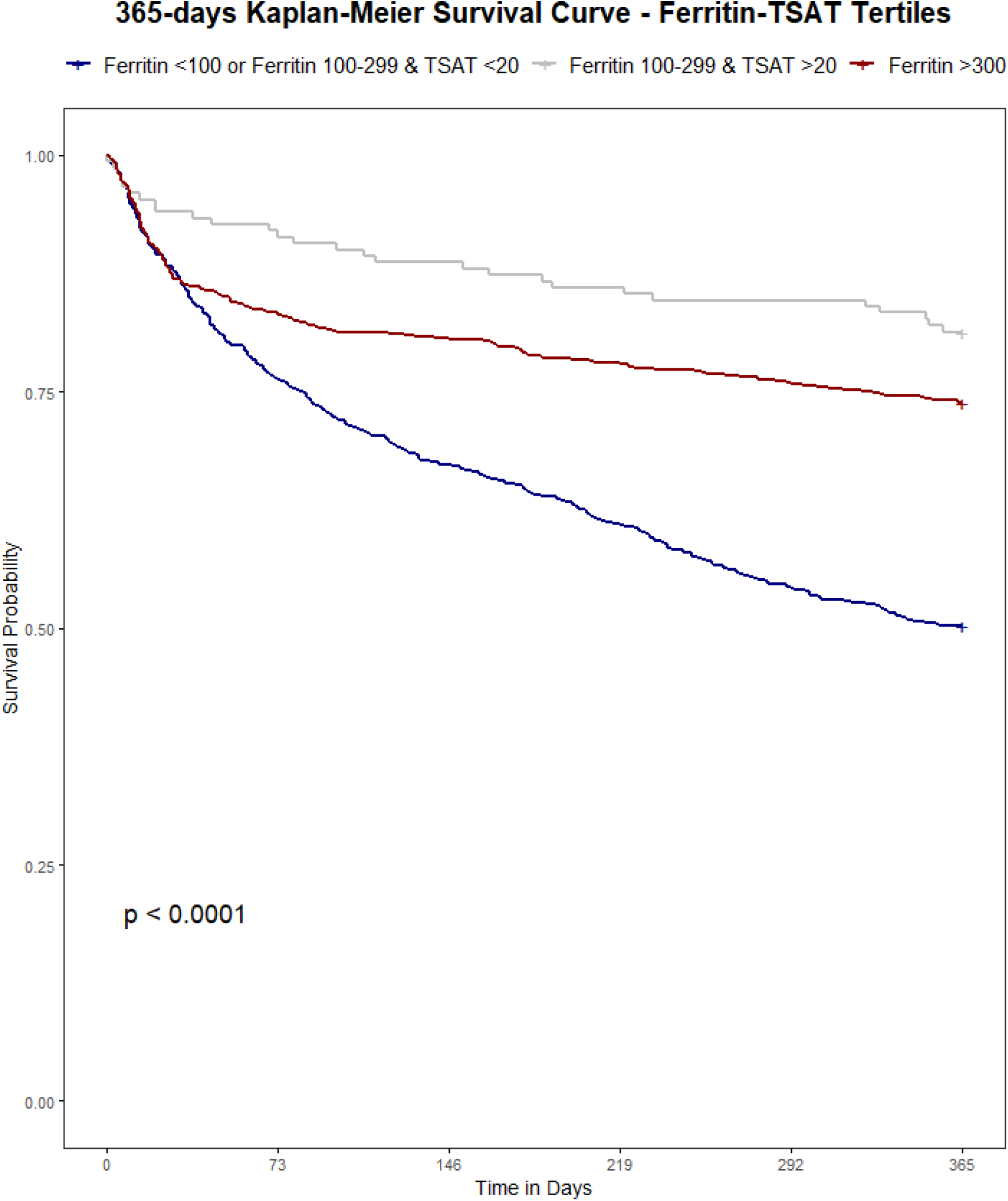
Kaplan–Meier survival to 365 days based on a combined Ferritin-TSAT category model. Population: hospitalized CHF index admissions. Groups: Deficient (ferritin <100 μg/L or 100–299 μg/L with TSAT <20%), Hyperferritinemia (ferritin ≥300 μg/L), Intermediate (ferritin 100–299 μg/L with TSAT ≥20%). Time zero: admission. Outcome: all-cause mortality to day 365. Deaths were ascertained from linked death records and counted within 365 days of admission, including events occurring after 2019; non-mortality follow-up was restricted to the dataset timeframe. Statistics: log-rank test with numbers at risk shown. Interpretive note: early and sustained curve separation is consistent with distinct deficiency vs. inflammation/congestion phenotypes. Abbreviations: TSAT, transferrin saturation.

**Table 2.** Hazard ratios and confidence intervals for iron deficiency regressions across multiple crude and adjusted multivariate and univariate models. Tertile category models encompass multiple values as they categorize data into three distinct groups and show reference categories: TSAT <10%; ferritin <100 μg/L; combined guideline-defined iron deficiency. HR < 1 indicates lower hazard vs. reference, not absolute benefit. categories 2 and 3 HRs are calculated in relation to Tertile 1, with each Tertile referencing categories as seen in Figures 1-4 and defined with each category model. Reported HRs are from Model 3 (fully adjusted) unless specified. Tables S1-S4 show full multivariate models. Abbreviations: CI, confidence interval; TSAT, transferrin saturation.

| Iron Variable<br>Model | Model 1 | Model 2 | Model 3 | Univariate |
| --- | --- | --- | --- | --- |
| Guidelines-based<br>Model | **3.350** (95%CI 2.730–4.110), $p<0.001$ | **3.787** (95%CI 3.053–4.697), $p<0.001$ | **4.356** (95%CI 3.349–5.338), $p<0.001$ | **2.48** (95%CI 2.09–2.94), $p<0.001$ |
| TSAT Category<br>Model | *Tertile 2: **0.753** (95%CI 0.610–0.930), $p=0.009$*<br>*Tertile 3: **0.609** (95%CI 0.484–0.766), $p<0.001$* | *Tertile 2: **0.780** (95%CI 0.626–0.971), $p=0.026$*<br>*Tertile 3: **0.625** (95%CI 0.492–0.794), $p<0.001$* | *Tertile 2: **0.804** (95%CI 0.644–1.003), $p=0.053$*<br>*Tertile 3: **0.632** (95%CI 0.494–0.810), $p<0.001$* | *Tertile 2: **0.72** (95%CI 0.613–0.849), $p<0.001$*<br>*Tertile 3: **0.584** (95%CI 0.424–0.783), $p<0.001$* |
| Ferritin Category<br>Model | *Tertile 2: **0.671** (95%CI 0.544–0.827), $p<0.001$*<br>*Tertile 3: **0.400** (95%CI 0.313–0.511), $p<0.001$* | *Tertile 2: **0.649** (95%CI 0.523–0.804), $p<0.001$*<br>*Tertile 3: **0.362** (95%CI 0.280–0.468), $p<0.001$* | *Tertile 2: **0.670** (95%CI 0.538–0.833), $p<0.001$*<br>*Tertile 3: **0.308** (95%CI 0.234–0.404), $p<0.001$* | *Tertile 2: **0.662** (95%CI 0.507–0.813), $p<0.001$*<br>*Tertile 3: **0.388** (95%CI 0.304–0.574), $p<0.001$* |
| Ferritin-TSAT<br>Category Model | *Tertile 2: **0.337** (95%CI 0.228–0.496), $p<0.001$*<br>*Tertile 3: **0.498** (95%CI 0.328–0.517), $p<0.001$* | *Tertile 2: **0.338** (95%CI 0.228–0.501), $p<0.001$*<br>*Tertile 3: **0.551** (95%CI 0.389–0.622), $p=0.001$* | *Tertile 2: **0.356** (95%CI 0.238–0.530), $p<0.001$*<br>*Tertile 3: **0.495** (95%CI 0.380–0.512), $p<0.001$* | *Tertile 2: **0.306** (95%CI 0.228–0.496), $p<0.001$*<br>*Tertile 3: **0.517** (95%CI 0.315–0.583), $p<0.001$* |

Model discrimination was satisfactory and subgroup analyses confirmed stable performance with minimal variation in hazard estimates within demographic and clinical strata (Table 3). The choice of 65 years as an age cutoff during subgroup analysis aligned with standard practice in cardiology research.

**Table 3.**
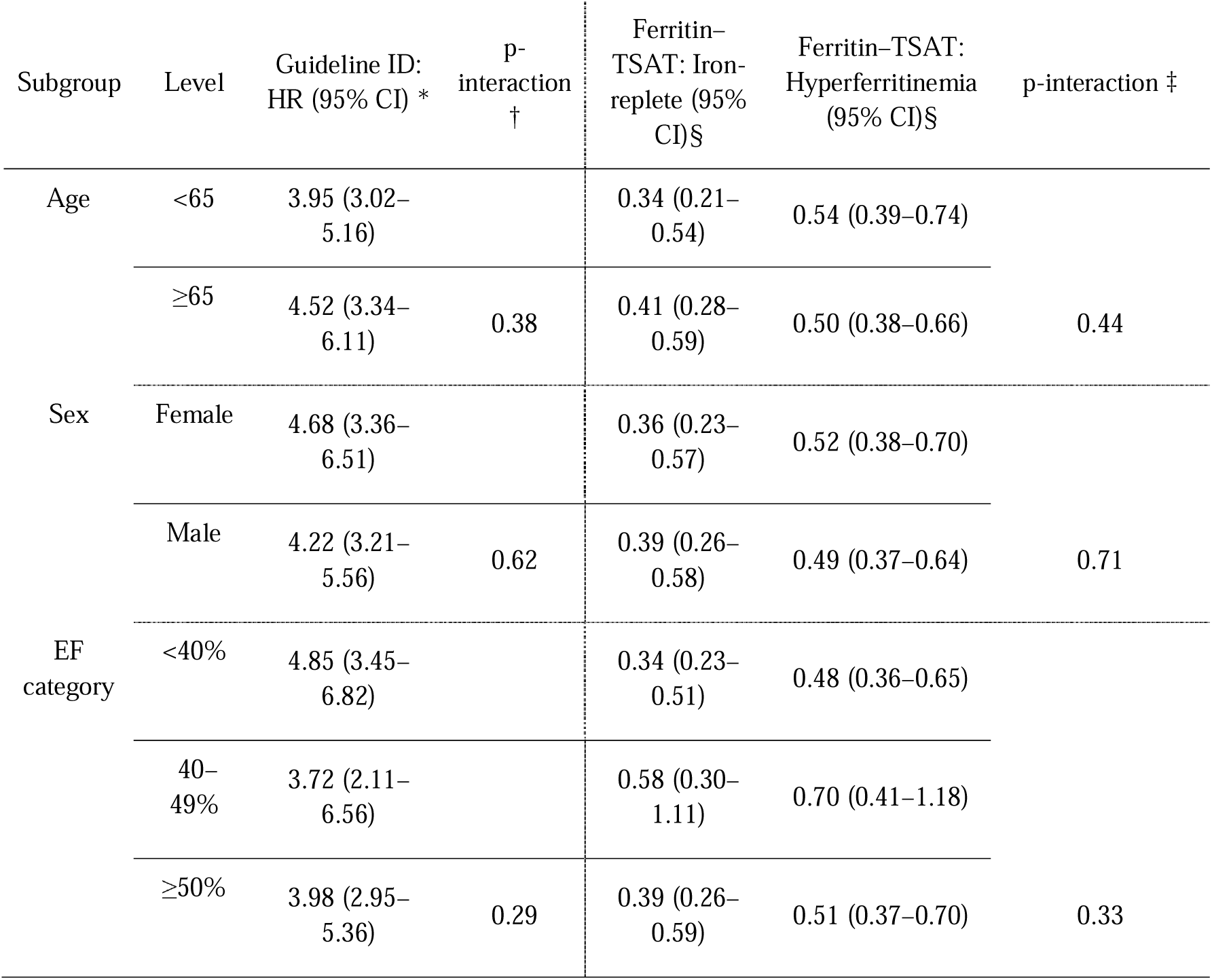
Subgroup analyses of 365-day mortality by iron status across prespecified strata. Values are hazard ratios (HRs) with 95% confidence intervals from fully adjusted Cox models. * Guideline ID compares ID vs non-ID within each stratum; § Ferritin–TSAT reference = True ID (ferritin <100 or 100–299 with TSAT <20%); † p-interaction (1-df) for Guideline ID × subgroup; ‡ p-interaction (2-df, global) for Ferritin–TSAT × subgroup. P-interaction was calculated using Wald test. Abbreviations: CI, confidence interval; df, degrees of freedom; EF, ejection fraction; HR, hazard ratio; ID, iron deficiency; TSAT, transferrin saturation.

## Discussion

This study demonstrates that iron dysregulation in CHF—spanning deficiency to hyperferritinemia—serves as a significant prognostic indicator of one-year survival. By evaluating novel categorizations of iron deficiency, we move beyond binary classifications to identify distinct risk phenotypes. Among the tested categorical frameworks, and as outlined in Table 2, the guideline-based definition provided superior risk stratification for adverse outcomes in high-risk patients during multivariate hazard analysis, while the combined Ferritin-TSAT Category Model revealed an iron store subgroup with markedly reduced mortality risk. As detailed in Tables 2 and S1–S4, the iron-status coefficients changed little with progressive adjustment, while clinical covariates behaved consistently (age, shock, malignancy with increased risk; CABG, ACEi/ARB, β-blocker, IV iron with decreased risk).

The prognostic implications of iron deficiency and hyperferritinemia in hospitalized CHF patients are profound; a study of 1506 CHF patients identified iron deficiency as a strong and independent predictor of mortality (HR 1.42, 95%CI 1.14–1.77, p = 0.002) [1]. A stark mortality gradient emerged between iron deficient and non-iron deficient cohorts (56.1% vs. 29.4% at one year p < 0.001), highlighting iron homeostasis as pivotal to cardiac and systemic function. In this cohort, guideline-defined iron deficiency was paradoxically associated with shorter hospital length of stay and lower rates of surgical and ICU admissions, despite substantially higher 1-year mortality compared with non-iron-deficient patients. These utilization patterns likely reflect differences in case-mix and admission type rather than genuinely lower overall risk: iron-deficient patients were older and predominantly managed medically, with fewer procedures and less ICU exposure, yet they experienced the worst long-term survival. By contrast, the non-iron-deficient group encompassed many hyperferritinemic patients (ferritin ≥300 µg/L), a phenotype that in other acute-illness cohorts is strongly linked to systemic inflammation, sepsis, multiorgan dysfunction, greater need for ICU care, and longer ICU and hospital stays. This suggests that hyperferritinemia in hospitalized CHF may mark a more acutely inflamed and clinically complex subgroup that requires prolonged hospitalization, even though their adjusted 365-day mortality in our data was lower than that of the most iron-deficient patients. Consistent with this pattern, the iron-deficient subgroup had a lower prevalence of AKI, paralleling their lower exposure to ICU care, mechanical ventilation, and vasopressors—factors that are well established to increase AKI risk in critically ill populations. Notably, Kaplan–Meier analyses underscored key distinctions among models (all p < 0.001): while guideline-based classifications exhibited the strongest survival divergence overall (Figure 1), they, alongside TSAT- and ferritin-only category models (Figures 2, 3), showed limited prognostic utility within the first 30 days. In contrast, the Ferritin-TSAT Category Model (Figure 4) demonstrated sustained late-term yet stronger early term stratification, distinguishing “true” non-iron deficiency patients, the iron-replete subgroup, from the two high-risk subgroups. This showed earlier and sustained curve separation, with speculative mechanisms involving early risks, reflecting impaired oxygen delivery/energetics, whereas late risks reflect persistent inflammatory or congestion-related iron sequestration [2, 24]. For reference, the iron deficiency cohort in the Ferritin-TSAT Category Model showed a consistent survival pattern due to its fixed, guideline-definition across these analyses (Figure 4).

Current guideline criteria, though clinically practical, conflate hyperferritinemic (≥300 µg/L) and normoferritinemic patients under a single "non-iron deficient" label, obscuring inflammation- or congestion-driven ferritin and iron elevations [10,18,16,25]. TSAT categories (<10%) effectively isolate severe functional deficiency but fail to discern elevated ferritin driven by said inflammation or congestion. Conversely, ferritin categories identify hyperferritinemia but overlook functional deficiency in patients with normal ferritin but critically low TSAT. The Ferritin-TSAT Category Model addresses these gaps by delineating three phenotypes: (1) true deficiency (low ferritin/TSAT), (2) intermediate-risk, hyperferritinemia, and (3) an iron-replete cohort. Studies show that iron-replete patients often fare better than hyperferritinemic status, substantiating the value of a combined biomarker approach for more precise patient stratification [16,18,24]. Notably, the combined Ferritin–TSAT model isolates a small iron-replete subgroup with the lowest risk, distinct from both the deficient and hyperferritinemic phenotypes (Tables S1–S4). This pattern strengthens the case for moving beyond a binary ‘deficient vs non-deficient’ framework when risk-stratifying hospitalized CHF patients.

ShortLterm prognostic results showed an immediate risk carried by severely deficient patients and some hyperferritinemic individuals, whereas long-term mortality confirmed that uncorrected iron dysregulation, be it deficiency or high ferritin, contributes to heart failure progression and mortality [26,27]. By unmasking the prognostic and therapeutic relevance of iron dysregulation subtypes, this model suggests limitations of the current binary paradigm and may inform future clinical guidelines, which currently overlook this maldistribution as a modifiable risk factor. Because nonLbinary criteria may help isolate hyperferritinemic individuals, clinicians could better allocate iron therapy, whether that therapy involves supplementation or chelation. Thus, the adoption of these new categorizations, in conjunction with other pertinent clinical and laboratory measures, may allow for a more individualized approach to preventing adverse outcomes in CHF. For instance, the ACC/AHA and ESC guidelines for CHF management emphasize iron deficiency correction but lack criteria to address hyperferritinemia or guide differential workups [28,26]. Integrating the Ferritin-TSAT Category Model into future iterations would enable tailored management, aligning therapies with underlying pathophysiology: intravenous iron for true deficiency, diuresis or anti-inflammatories for congestion- or inflammation-mediated hyperferritinemia, and close monitoring for intermediate-risk patients [16,29]. Although IV iron, ACEi/ARB and β-blockers were associated with lower hazard after adjustment, these findings remain hypothesis-generating due to potential residual confounding and treatment-selection bias.

Recent large-scale datasets have refined how iron deficiency is defined in heart failure and frame our inpatient findings. In the Swedish Heart Failure Registry, compared ESC guideline criteria, IRONMAN trial criteria, TSAT <20%, and ferritin <100 μg/L across >20,000 mainly ambulatory patients with HFrEF, HFmrEF, and HFpEF. Iron deficiency was common and consistently linked to worse symptoms and poorer quality of life, but TSAT <20% and IRONMAN criteria showed the clearest associations with mortality and heart failure hospitalizations, whereas ferritin <100 μg/L alone carried limited prognostic value [30]. These outpatient data support our focus on TSAT-based and combined ferritin–TSAT definitions as markers of clinically relevant iron deficiency. In contrast, guideline-defined iron deficiency in our acutely hospitalized cohort was associated with 365-day all-cause mortality, implying that identical biochemical thresholds identify a more adverse phenotype once decompensation occurs. The IRONMAN trial extends this prognostic information to treatment response. In 1,137 outpatients with chronic heart failure (EF ≤45%) and iron deficiency (ferritin <100 μg/L or TSAT <20%), intravenous ferric derisomaltose produced larger hemoglobin gains and greater absolute reductions in heart failure hospitalization or cardiovascular death in patients with lower baseline TSAT and more severe anemia, whereas those with TSAT ≥20% or without anemia had few events and little benefit [31]. These findings mirror our observation that the most severely iron-deficient inpatients experience the greatest excess mortality and are likely to derive the largest absolute benefit from targeted iron repletion. The EDIFICA acute heart failure cohort complements these results by dissecting ferritin as a marker of both iron status and inflammation. In 526 patients, higher admission ferritin independently predicted a higher 180-day risk of heart failure hospitalization or cardiovascular death, whereas low ferritin and guideline-defined iron deficiency were not robust markers of risk; at discharge, defective iron utilization or guideline-defined iron deficiency were paradoxically associated with lower risk than apparent iron repletion [32]. EDIFICA therefore concludes that hyperferritinemia in acute heart failure frequently reflects an inflammatory “hyperferritinemic” phenotype with restricted iron bioavailability and that reliance on ferritin thresholds alone risks undertreating this subgroup: an interpretation that aligns with our separation of a hyperferritinemic inpatient phenotype, thereby supporting a more nuanced, phenotype-oriented approach than a simple binary “deficient versus non-deficient” framework, and our emphasis on TSAT and combined ferritin–TSAT frameworks for risk stratification.

Early iron parameter assessment (within 24 hours of admission) could identify high-risk patients prone to early decompensation, as both deficiency and hyperferritinemia correlate with rapid clinical decline. Multivariate analyses confirmed iron dysregulation’s independent mortality risk, unaffected by covariates such as shock, malignancy, or ICU admission. Importantly, standard CHF therapies (beta-blockers, ACE inhibitors/ARBs) synergize with iron status optimization. In patients with low ferritin or TSAT, especially below the accepted guideline thresholds, intravenous iron therapy appears to confer significant survival benefits, with hazard ratios suggesting protective effects [14]. Early repletion reduces 14–28-day mortality and may attenuate one-year risk, whereas hyperferritinemic patients, often misclassified as "non-deficient," risk undertreatment.

## Limitations

Several limitations warrant caution in interpreting these findings. The dataset is based on a retrospective review of records from a single center, Beth Israel Deaconess Medical Center, which may limit generalizability to other institutions or healthcare settings with different patient demographics and clinical practices. Outcomes beyond all-cause mortality could not be reliably captured; it is the only reliable outcome to extract from this study using the single-center dataset. Mortality can be verified post-discharge and in other centers and is a major outcome. As with most retrospective analyses, causal inferences cannot be definitively established, and observed associations between iron parameters and outcomes may be influenced by unmeasured confounders. During dataset creation, some inflammatory markers were not routinely measured, making it difficult to distinguish between true iron overload (e.g., hemochromatosis) from inflammation-driven ferritin elevations due to limited etiologic coding and inflammatory biomarkers; future work incorporating CRP, hepcidin, and sTfR is warranted. Inflammation was partially assessed, as neutrophil count was initially measured, but interpreted to have no significant influence. Addressing this gap in future investigations could provide a clearer understanding of inflammatory mechanisms in iron dysregulation and heart failure progression.

### Future Directions

The need for prospective validation of these findings should be emphasized, stratifying patients into nuanced ferritin–TSAT categories and testing personalized iron therapies. The analysis includes large numbers of patients and uses multiple adjustment models that incorporate age, shock, malignancy, renal function, ejection fraction, and comedications, bolstering that iron status itself is an independent risk factor. By comparing guideline-defined iron deficiency, TSAT-based regression, ferritin-based categories, and a combined ferritin–TSAT model, the study gives a comprehensive perspective on how best to capture diverse risk groups in CHF. However, incorporating dedicated inflammatory biomarkers (CRP, hepcidin, soluble transferrin receptor [24]) would clarify whether hyperferritinemia is truly synonymous with iron repletion or driven by an acute-phase response that blocks normal iron utilization. Tracking iron indices beyond a single time point may also show how dynamic changes in iron status correlate with readmissions or advanced interventions for CHF. In turn, guidelines committees might adopt broader screening criteria that identify not only classical deficiency but also potential hyperferritinemic or inflammatory states.

## Conclusion

This study reaffirms iron status—both extremely deficient and excessively high—as a central determinant of CHF outcomes. While guideline-defined iron deficiency remains a strong, practical predictor of early and longer-term mortality, a significant proportion of patients with ferritin ≥300 μg/L may also exhibit poor survival trajectories. Non-binary models offer finer discrimination between true deficiency and hyperferritinemia, guiding either supplementation or further investigation of systemic drivers. Tailoring interventions based on iron parameters, such as prioritizing IV iron in deficiency or investigating inflammation in hyperferritinemia, exemplifies a precision-medicine approach to heart failure. By identifying iron misregulation early in the course of hospitalization, clinicians may optimize IV iron use, investigate inflammatory pathologies, and improve overall survival for patients grappling with CHF, although further trials are needed to define optimal timing and protocols.

## Author Contributions

Conceptualization, A.H..; Methodology, A.H.; Formal Analysis, A.H..; Data Curation, A.H..; Writing—Original Draft Preparation, A.H., A.S., S.D.; Writing—Review and Editing, A.S., A.H.; Visualization, A.S., A.H.

## Institutional Review Board Statement

Ethical review and approval were waived for this study due to the use of fully de-identified data from the MIMIC-IV database, in accordance with the policies of the Institutional Review Boards of the Massachusetts Institute of Technology and Beth Israel Deaconess Medical Center.

## Informed Consent Statement

Patient consent was waived due to the use of de-identified retrospective data.

## Data Availability Statement

The data that support the findings of this study are derived from the MIMIC-IV database, which is publicly available at https://physionet.org/content/mimiciv/2.2/ for credentialed researchers who complete the required data use agreement.

## Supplementary Materials

The following supporting information can be downloaded: Table S1–S4.

---

**Table S1.** Fully adjusted hazard ratios and confidence intervals for covariates with the Guidelines-based Model. Abbreviations: ICU: Intensive care unit; eGFR: estimated glomerular filtration rate; CABG: coronary artery bypass graft; PCI: percutaneous coronary intervention; ACEi/ARB: Angiotensin converting enzyme inhibitor/Angiotensin II receptor blockers; IV: intravenous. Harrell’s C-index 0.72 (0.70-0.74); AIC 1650.21; Brier 0.20.

|  | HR | CI |
| --- | --- | --- |
| Age | 1.036 | 95% CI 1.03-1.05, p<0.001 |
| Sex | 1.105 | 95% CI 0.92-1.34, p=0.299 |
| Mildly-Reduced Ejection Fraction | 0.788 | 95% CI 0.51-1.24, p=0.299 |
| Preserved Ejection Fraction | 0.887 | 95% CI 0.74-1.08, p=0.212 |
| ICU Stay | 1.480 | 95% CI 1.19-1.86, p=0.001 |
| Acute Coronary Artery Disease | 2.126 | 95% CI 1.18-3.84, p=0.012 |
| Hypertension | 1.130 | 95% CI 0.87-1.48, p=0.374 |
| Shock | 1.978 | 95% CI 1.53-2.56, p<0.001 |
| Diabetes | 1.496 | 95% CI 1.24-1.81, p<0.001 |
| Malignant Cancer | 1.501 | 95% CI 1.14-1.98, p=0.004 |
| Hemoglobin (log10) (g/dL) | 0.687 | 95% CI 0.25-1.95, p=0.48 |
| eGFR (log10) (mL/min/1.73m<sup>2</sup>) | 0.738 | 95% CI 0.52-1.07, p=0.101 |
| CABG | 0.314 | 95% CI 0.14-0.72, p=0.005 |
| PCI | 0.653 | 95% CI 0.40-1.08, p=0.097 |
| ACEi/ARB Use | 0.684 | 95% CI 0.56-0.84, p<0.001 |
| Dialysis Status | 0.471 | 95% CI 0.33-0.68, p<0.001 |
| IV Iron Supplementation | 0.571 | 95% CI 0.45-0.73, p<0.001 |
| Beta-Blocker Use | 0.659 | 95% CI 0.53-0.82, p<0.001 |
| Calcium Channel Blocker Use | 0.673 | 95% CI 0.53-0.87, p=0.002 |

**Table S2.** Fully adjusted hazard ratios and confidence intervals for covariates with the TSAT Category Model. Abbreviations: ICU: Intensive care unit; eGFR: estimated glomerular filtration rate; CABG: coronary artery bypass graft; PCI: percutaneous coronary intervention; ACEi/ARB: Angiotensin converting enzyme inhibitor/Angiotensin II receptor blockers; IV: intravenous. Harrell’s C-index 0.71 (0.68-0.74); AIC 1680.84; Brier 0.23.

|  | HR | CI |
| --- | --- | --- |
| Age | 1.045 | 95%CI 1.04-1.05, p<0.001 |
| Sex | 1.275 | 95%CI 0.99-1.44, p=0.078 |
| Mildly-Reduced Ejection Fraction | 0.699 | 95%CI 0.50-1.23, p=0.278 |
| Preserved Ejection Fraction | 0.775 | 95%CI 0.67-0.99, p=0.031 |
| ICU Stay | 1.247 | 95%CI 1.15-1.80, p=0.002 |
| Acute Coronary Artery Disease | 1.579 | 95%CI 0.85-2.75, p=0.159 |
| Hypertension | 1.179 | 95%CI 0.89-1.52, p=0.280 |
| Shock | 1.836 | 95%CI 1.40-2.34, p<0.001 |
| Diabetes | 1.560 | 95%CI 1.25-1.82, p<0.001 |
| Malignant Cancer | 1.398 | 95%CI 1.12-1.95, p=0.006 |
| Hemoglobin (log10) (g/dL) | 0.506 | 95%CI 0.21-1.72, p=0.340 |
| eGFR (log10) (mL/min/1.73m<sup>2</sup>) | 1.046 | 95%CI 0.55-1.14, p=0.209 |
| CABG | 0.255 | 95%CI 0.11-0.53, p<0.001 |
| PCI | 0.685 | 95%CI 0.37-1.00, p=0.050 |
| ACEi/ARB Use | 0.685 | 95%CI 0.54-0.82, p<0.001 |
| Dialysis Status | 0.564 | 95%CI 0.34-0.69, p<0.001 |
| IV Iron Supplementation | 0.825 | 95%CI 0.54-0.87, p=0.002 |
| Beta-Blocker Use | 0.681 | 95%CI 0.53-0.82, p<0.001 |
| Calcium Channel Blocker Use | 0.684 | 95%CI 0.53-0.87, p=0.002 |

**Table S3.** Fully adjusted hazard ratios and confidence intervals for covariates with the Ferritin Category Model. Abbreviations: ICU: Intensive care unit; eGFR: estimated glomerular filtration rate; CABG: coronary artery bypass graft; PCI: percutaneous coronary intervention; ACEi/ARB: Angiotensin converting enzyme inhibitor/Angiotensin II receptor blockers; IV: intravenous. Harrell’s C-index 0.73 (0.70-0.75); AIC 1610.19; Brier 0.18

|  | HR | CI |
| --- | --- | --- |
| Age | 1.040 | 95% CI 1.04-1.05, p<0.001 |
| Sex | 1.186 | 95% CI 0.99-1.44, p=0.078 |
| Mildly-Reduced Ejection Fraction | 0.780 | 95% CI 0.50-1.23, p=0.278 |
| Preserved Ejection Fraction | 0.811 | 95% CI 0.67-0.99, p=0.031 |
| ICU Stay | 1.433 | 95% CI 1.15-1.80, p=0.002 |
| Acute Coronary Artery Disease | 1.526 | 95% CI 0.85-2.75, p=0.159 |
| Hypertension | 1.160 | 95% CI 0.89-1.52, p=0.280 |
| Shock | 1.805 | 95% CI 1.40-2.34, p<0.001 |
| Diabetes | 1.507 | 95% CI 1.25-1.82, p<0.001 |
| Malignant Cancer | 1.472 | 95% CI 1.12-1.95, p=0.006 |
| Hemoglobin (log10) (g/dL) | 0.599 | 95% CI 0.21-1.72, p=0.340 |
| eGFR (log10) (mL/min/1.73m<sup>2</sup>) | 0.792 | 95% CI 0.55-1.14, p=0.209 |
| CABG | 0.231 | 95% CI 0.11-0.53, p<0.001 |
| PCI | 0.604 | 95% CI 0.37-1.00, p=0.050 |
| ACEi/ARB Use | 0.661 | 95% CI 0.54-0.82, p<0.001 |
| Dialysis Status | 0.477 | 95% CI 0.34-0.69, p<0.001 |
| IV Iron Supplementation | 0.681 | 95% CI 0.54-0.87, p=0.002 |
| Beta-Blocker Use | 0.658 | 95% CI 0.53-0.82, p<0.001 |
| Calcium Channel Blocker Use | 0.674 | 95% CI 0.53-0.87, p=0.002 |

**Table S4.** Fully adjusted hazard ratios and confidence intervals for covariates with the Ferritin-TSAT Category Model. Abbreviations: ICU: Intensive care unit; eGFR: estimated glomerular filtration rate; CABG: coronary artery bypass graft; PCI: percutaneous coronary intervention; ACEi/ARB: Angiotensin converting enzyme inhibitor/Angiotensin II receptor blockers; IV: intravenous. Harrell’s C-index 0.76 (0.73-0.78); AIC 1580.07; Brier 0.16

|  | HR | CI |
| --- | --- | --- |
| Age | 1.039 | 95% CI 1.03-1.05, p<0.001 |
| Sex | 1.191 | 95% CI 0.99-1.44, p=0.070 |
| Mildly-Reduced Ejection Fraction | 0.774 | 95% CI 0.50-1.22, p=0.262 |
| Preserved Ejection Fraction | 0.812 | 95% CI 0.68-0.99, p=0.031 |
| ICU Stay | 1.437 | 95% CI 1.15-1.80, p=0.002 |
| Acute Coronary Artery Disease | 1.922 | 95% CI 1.06-3.50, p=0.032 |
| Hypertension | 1.163 | 95% CI 0.89-1.53, p=0.271 |
| Shock | 1.847 | 95% CI 1.43-2.40, p<0.001 |
| Diabetes | 1.488 | 95% CI 1.24-1.80, p<0.001 |
| Malignant Cancer | 1.474 | 95% CI 1.12-1.95, p=0.006 |
| Hemoglobin (log10) (g/dL) | 0.599 | 95% CI 0.21-1.72, p=0.340 |
| eGFR (log10) (mL/min/1.73m<sup>2</sup>) | 0.824 | 95% CI 0.58-1.19, p=0.295 |
| CABG | 0.224 | 95% CI 0.10-0.51, p<0.001 |
| PCI | 0.646 | 95% CI 0.39-1.08, p=0.090 |
| ACEi/ARB Use | 0.683 | 95% CI 0.56-0.84, p<0.001 |
| Dialysis Status | 0.505 | 95% CI 0.36-0.73, p<0.001 |
| IV Iron Supplementation | 0.670 | 95% CI 0.53-0.86, p=0.001 |
| Beta-Blocker Use | 0.665 | 95% CI 0.54-0.83, p<0.001 |
| Calcium Channel Blocker Use | 0.682 | 95% CI 0.53-0.88, p=0.003 |

